# Trends and Determinants of Neonatal Mortality in Rural Ethiopia

**DOI:** 10.1101/2024.03.18.24304498

**Authors:** Sintayehu Asaye, Dawit Saketa, Dires Birhanu, Tadesse Gudeta, Merga Besho, Masrie Getnet, Gurmesa Tura Debelew, Negalign Berhanu, Yibeltal Siraneh, Fira Abamecha, Dessalegn Tamiru

**Affiliations:** Institute of Health, Jimma University, Jimma, Ethiopia; College of Natural and Computational Sciences, Wollega University, Wollega, Ethiopia; College of Health Science, Dilla University, Dilla, Ethiopia; Institute of Health Science, Wollega University, Wollega, Ethiopia

**Author notes:** Correspondence: Sintayehu Asaye (P.O. Box 378, Jimma University, Ethiopia).

**Keywords:** Neonatal mortality, trends, Determinants, Rural, Ethiopia

## Abstract

**Background:** Neonatal mortality is a significant challenge that affects babies within the first 28 days of life. The issue is particularly challenging for healthcare systems in developing countries, where interventions are required. Although there has been a decline in neonatal mortality worldwide, comprehensive data on the patterns of neonatal mortality and the contributing factors in rural regions of Ethiopia is lacking.

**Objective:** To determine neonatal mortality trends and mortality in rural Ethiopia using 2011-2019 DHS data

**Methods:** Ethiopian demographic health survey (EDHS) program conducted a cross-sectional community-based study in rural Ethiopia in 2011, 2016, and 2019. The study included women who gave birth within the specified timeframe and agreed to participate. Sampling was done through a multistage cluster approach, and STATA version 17 was used to analyze the data. Predictor variables were validated through multiple logistic regression analysis. Weighted estimates were used to derive population-level statistics and a p-value less than 0.05 was considered significant.

**Results:** The study analyzed data from 22,755 women who participated in EDHS surveys between 2011 and 2019. Neonatal mortality rates decreased from 7.5% to 6.03%. Regional variations were observed, with Gambela and Tigrai having the lowest rates, and Dire Dawa and the Somali region having higher rates. Factors like mother’s age, wealth index, birth order, neonate’s sex, and presence of twins, immediate breastfeeding, and baby’s size were associated with neonatal mortality.

**Conclusion and recommendation:** Despite significant advancements that have been made to decrease neonatal mortality, there remain challenges that need to be addressed. Therefore, regional health bureaus should strengthen their strategies to enhance antenatal care (ANC) visits and promote birth delivery at health facilities.

## Background

Neonates are newborns up to 28 days old. [1,2]. Neonates can be classified by gestational age, birth weight, and based on the combination of gestational age with birth weight[3].

Neonatal death is a serious problem that occurs within the first 28 days of life due to different factors which can be from prenatal, intra-natal, or post natal driven causes [4,5] .Commonly, neonatal mortality is expressed by the rate in which it states that the number of deaths during the first four weeks of life per 1000 live births in a given time. Based on time to death, neonatal mortality can be subdivided into very early (<1 day), early (1-7days), and late neonatal deaths [6].

Neonatal mortality rate is the highest within the first seven days of life, with 75% of deaths occurring during that time. Roughly one million neonates die within 24 hours of being born. Preterm-related complications, perinatal-related complications, and infections are the leading causes of neonatal mortality.[6].

The three major causes of neonatal deaths globally are infections (36%), pre-term delivery (28%), and birth asphyxia (23%) [7]. Africa loses 1.12 million neonates yearly due to preventable causes including prematurity, low birth weight, infections, lack of oxygen, and birth trauma. [8].

In 2021, 2.3 million children died in the first month of life worldwide or approximately 6,400 newborns per day. The annual death toll could reach up to 2.5 million people, mostly in low- and middle-income countries (LMICs), where basic human rights are completely ignored[9,10]. Of the total global neonatal mortality, 80% took place in Sub-Saharan Africa and Southern Asia, of which two-thirds and three-quarters of the deaths occurred on the first and seventh day of life respectively. The place of neonatal birth matters the survival which increases by 20 times higher in sub-Saharan Africa and Southern Asia and higher for newborns during conflicts[9].

The burden of neonatal mortality is not only limited to the neonates themselves but has multilevel effects on country profile, family spiritual life, family health, family social relation, family economy, and siblings and it can cause divorce to the partner due to the blame, mistreated, and dishonored[11,12].Neonatal mortality is painful full which can cause for parental psychosocial problems which may result in self-suicide to the family[11].

Professionals who work in neonatal intensive care units are more likely to be present during the end-of-life time for neonates, which can hurt their emotions and practice. Coping with the death of newborns is a highly stressful task that can lead to post-traumatic stress symptoms and burnout in professionals[13].

The Sustainable Development Goals (SDG) aims to end preventable deaths of newborns and children under the age of five by 2030. All countries should strive to reduce newborn mortality to at least 12 deaths per 1000 live births and under-five mortality to at least 25 deaths per 1000 live births. However, in rural Ethiopia, the neonatal mortality rate is still very high, standing at 62 deaths per 1000 live births. This is significantly higher than the SDG targets[14].

As per current trends of neonatal mortality, it is projected that 1·8 million neonates would die in 2030, however if each country achieved the SDG target, there would be about 1·2 million neonatal deaths. Achieving this goal requires increased efforts to enable continued improvements in child survival[15].

The study conducted in China showed the neonatal mortality had steadily decreased from 5.9 deaths per 1000 live births in 2014 to 3.9 deaths per 1000 live births in 2018, which was lower than the target of SDG3 at the national level[16].The systematic review conducted in Brazil showed that the average neonatal mortality rate was 9.46 deaths/1,000 live births in 2017, with a reduction of 2.15% between 2007 to 2017[17].In the systematic review from South East Asia Countries Indonesia was the country with the highest neonatal mortality of 61 live births per 1000 live birth in 2017, the trend decreased gradually from 103 per 1000 live birth in 2000[18]. In Nepal though the neonatal mortality is decreasing, if it continues with current rate, it will take another 50 years for the country to attain the 2030 SDG target. NMR declined by an average of only 4.0% between 2001 and 2016[19].In Ghana there has been a steady decline in neonatal mortality from 30 per 1000 live birth in 1998 to 25 per 1000 live births in 2017 for all births [20]. In Burundi, the NMR was declined from 38.7 per 1000 live births in 2010 to 25.0 deaths per 1000 live births in 2016, respectively. The richer wealth quintile has slightly higher NMR than other parts of the country[21].The review results conducted in Kenya showed that the neonatal mortality rate has fallen gradually from 35.4 deaths per 1000 live births in 1975 to 19.6 deaths per 1000 live births in 2018[8].

In Ethiopian the neonatal mortality rate declined by 1.9% per annum from 1995 to 2010, logarithmically[7]. However, neonatal death rates climbed from 29 per 1,000 live births in the Ethiopian demographic health survey (EDHS2016) to 30 per 1,000 live births in the Ethiopian mini demographic health survey (EMDHS 2019) report[7,21–23]. Despite its increment there are significant variations across specific regions of the country. For instance, in the Somalia region, around 130 newborns do not survive beyond the first week of life, the estimated NMR is 54 per 1,000 live births in Oromia, and 46 per 1000 live birth in Amhara region. Ethiopia’s progress was slow, and it fell well short of the UN’s ambitious aim of avoiding unnecessary infant deaths and lowering neonatal mortality to 12 per 1,000 live births in every nation by 2030[22].

It has been observed that the male neonate had a higher likelihood of dying in the first four week of life when compared to their female counterpart. This finding was consistently found in the study conducted using DHS data in Zambia[24], Afghanistan[25], Nigeria[26], Burundi[21] and Bangladesh[27].

Previous studies have established that prolongation of the birth interval between two subsequent pregnancies of more than 24 months (about 2 years) had a positive significant effect on neonatal survival. In the study conducted using DHS data in Nepal a birth interval shorter than 24 months (about 2 years) was found a significant predictor of neonatal mortality[20].In Zambia, the study from ZDHS data found that longer birth interval was associated with low neonatal mortality. Similarly, the study conducted in Ethiopia using EDHS 2016 data found a Children delivered at a birth interval of greater than 24 months (about 2 years) had higher chance of surviving their neonatal period[20,25].

In medical literature multiple pregnancy are classified as high-risk birth because they are associated with increased fetal and neonatal complications compared to singletons. This finding was similar in the study conducted using DHS in Ghana [20] and Afghanistan [25] and Ethiopia[7]. In the study conducted in Afghanistan the two extreme maternal age (age below 18 years and above 35 years old) are more likely linked to increased neonatal mortality when compared to maternal age between 20-30 years old[25].

In the retrospective cohort study conducted in Brazil[17] the neonate born to mothers who were having a child for the first time had higher chances of dying compared to those with mothers who were multiparous. Contradicting to this finding the DHS data study conducted in Ghana found that high parity was associated with a higher neonatal mortality[20].

In study conducted with DHS data from Ghana the risk of neonatal mortality Among mothers who did not attend ANC, was approximately five times as high as the risk of neonatal deaths for mothers who attended ANC. Consistent finding was reported across the study conducted in Afghanistan, Nigeria and Ethiopia[7,25,26].

Study conducted in Zambia also reveal that the survival chance for the children born to mother’s using contraceptives were 66 per cent more likely to compare to children born to mothers not using contraceptives[28]. Consistent to the study mentioned above, In the longitudinal study done in North-western Ethiopia mothers who were using contraceptives were two times less likely to lose a child compared to mothers who were not using contraceptives[20].

Neonatal mortality rates are still a significant problem and require ongoing research to address. Specifically, there is a need to focus on trends in rural areas of Ethiopia where there may be a lack of access to specialized facilities and professionals. While there have been some studies conducted in these areas, there is no evidence available to fully understand the trends in neonatal mortality rates and associated factors in rural areas of Ethiopia from 2011 to 2019. Therefore, this study aimed to analyze EDHS data from 2011 to 2019 to better understand these trends and contributing factors in rural Ethiopia.

## Materials and Methods

### Study Area, Design, and Period

Community-based cross-sectional study was conducted on the rural population across all administrative regions of Ethiopia by EDHS program from the period 2011, 2016, and 2019.

### Participant’s Selection and Exclusion

Regarding inclusion and exclusion criteria, these criteria are essential for defining the study population and ensuring that the analysis focuses on relevant data. In the context of neonatal mortality analysis using EDHS data:

#### Inclusion Criteria

- Participants within the specified age range
- Participants from the selected geographic areas covered by the EDHS.
- Participants for whom complete and relevant data on neonatal mortality and potential determinants are available.

#### Exclusion Criteria

- Participants outside the defined age range.
- Incomplete or missing data on neonatal mortality or key determinants.
- Participants from areas not covered by the EDHS.

### Source Population

All women of reproductive age living in rural Ethiopia during the EDHS surveys of 2011, 2016, and 2019 were considered as the source population.

### Study population

All pregnant women residing in rural areas of Ethiopia during 2011, 2016, and 2019, as recorded in the Ethiopian Demographic and Health Surveys (EDHS), were included as the study population.

### Sample Size Determination and Sampling Techniques

All pregnant who gave birth in rural Ethiopia during the years 2011, 2016, and 2019 were included in the Ethiopian Demographic and Health Surveys (EDHS) and households were selected via multistage cluster sampling to include pregnant women who gave birth and volunteered to participate in the study.

### Study variables

Neonatal mortality is defined as the death of a newborn within the first 28 days of life, as reported by the Ethiopian Demographic and Health Survey (EDHS). It is considered a dependent variable in this study. The independent variables that were considered in the study were age, region, altitude, religion, education, marital status, household access to media, source of drinking water, type of toilet facility, type of cooking fuel, wealth index of the household, number of living children, birth order number, number of antenatal visits, whether the child is a twin, sex of the child when it is put to the breast, place of delivery, delivery by cesarean section, and size of the child at birth.

### Data Quality Assurance

The data was extracted from the EDHS website after getting formal permission to access the data for research purposes only. Then the required data is meticulously exploited for the analysis and it was analyzed carefully using software and the data were also checked for missing values.

### Data Processing and Analysis

The neonatal mortality data from the DHS are analyzed using frequencies, percentages, cross tabulations, and multiple logistic regression statistical methods, taking into account the complex survey design. Weighted estimates are produced to generate population-level statistics.

Multiple logistic regression analysis was utilized where neonatal mortality coded as 0 and 1, where 0 indicates that the outcome of interest died, and 1 indicates absence outcome of interest, alive.

After a result from the logistic regression model with a logit link function, computing a marginal effect or risk difference often has numerous advantages as it estimates percentage point change, not a percent change. As we add more variables to the model, the marginal effects are not sensitive, and they remain unchanged. To compute the marginal effects, the change in probability when the discrete value of 1 of the independent variable changed is calculated.

Among the various logistic regression model fit assessment criteria, the commonly used measures, such as the Hosmer-Lemeshow test, the link test, the receiver operating characteristic (ROC) curve, pseudo R-squared, Residual Analysis, and outlier detection were used. All analyses were performed with Stata v. 17.

### Data Collection Techniques

Data were extracted from Ethiopia Demographic and Health Surveys (EDHS) conducted in 2011, 2016, and 2019 based on the outcome of interest, neonatal mortality. Request to EDHS made on Jan 15, 2024 to access the data for research purpose and we obtained the data on date Jan 16, 2024 with affirmation letter from EDHS team. The DHS employed a multistage cluster sampling design to select households for participation. The authors got a permission to access the EDHS data from the ICF-DHS program[29].

## Results

### Background characteristics

Out of the 22,755 participants, the majority of data were obtained from the 2011 EDHS: 9663 (42.5%), while 8667 (38.1%) were extracted from the 2016 EDHS. Many participating neonates, 9699 (42.6%), were obtained from an altitude between 1501 and 2300 meters high, followed by neonates from an altitude of 501 to 1500 meters high, 7515 (33%). That means more than 75% of the EDHS data on neonatal mortality used for this study were obtained from a cluster at an altitude of 501 to 2300 meters. Geographically, the DHS data used for this study were extracted from the rural distribution of all regions. Oromiya (16.4%), SNNPR(Southern Nations Nationalities Peoples’ Region) (14.5%), Amhara (11.2%), Afar (11.0%), and Somali (10.9%) regions tend to have a higher number of neonate participants in the study compared to the other rural regions. A smaller number of neonatal deaths in rural areas of Dire Dawa city administration and Harari region were included (4.1%, and 4.8%, respectively) **(Table 1).**

**Table 1:** Demographic Variables for trends and determinants of neonatal mortality in rural Ethiopia, EDHS, 2011, 2016 and 2019.

| Variable | Category | Frequency | Percentage |
| --- | --- | --- | --- |
| Ethiopian DHS Year | 2011 | 9663 | 42.5 |
|  | 2016 | 8667 | 38.1 |
|  | 2019 | 4425 | 19.4 |
| Cluster altitude in meters | <500m | 2337 | 10.3 |
|  | 501–1500m | 7515 | 33.0 |
|  | 1501–2300m | 9699 | 42.6 |
|  | >2300m | 3204 | 14.1 |
| Region | Tigray | 2,302 | 10.12 |
|  | Afar | 2503 | 11.0 |
|  | Amhara | 2538 | 11.2 |
|  | Oromiya | 3743 | 16.4 |
|  | Somali | 2487 | 10.9 |
|  | Benishangul-Gumuz | 2227 | 9.8 |
|  | SNNP | 3310 | 14.5 |
|  | Gambela | 1641 | 7.2 |
|  | Harari | 1082 | 4.8 |
|  | Dire Dawa | 922 | 4.1 |
| **Total** |  | **22755** | **100** |

Among 15623 (68.7%) of mothers have three or more living children, and 7132 (31.3%) have fewer than three living children. The age distribution of the mothers was examined, revealing a predominantly adult demographic profile. The majority of individuals fell within the 25–34 age range, constituting 11599 (51%) of the total population. The income distribution analysis demonstrated significant heterogeneity among the participating mothers. The majority of respondent mothers, 14050 (61.7%), indicated a poor wealth index, while 4683 (20.6%) and 4022 (17.7%) of mothers reported a rich and middle wealth index, respectively. The number of antenatal care service visits during pregnancy disparities was also observed among mothers.

While 50% of the women did not attain the optimal number of ANC service visits—at least four ANC visits—it has been reported that the attendance status of more than one-third of the mothers, 34.5%, was not reported. Only 15.4% of the mothers were reported to have attained the optimal level of at least four ANC visits. This suggests that a significant proportion of mothers in rural Ethiopia didn’t attend the optimal ANC visits, which may have implications for neonatal mortality.

The marital status of mothers indicates that the majorities of them (94.2%) are married and live with their husbands. Examining educational attainment levels reveals that a substantial portion of the participating mothers (72.0%) had no formal education. Educated cohorts tend to have higher rates of infant treatment, indicating improvements in neonatal mortality. The distribution of religious affiliations of mothers in the study was analyzed, and Muslims were found to be the substantial group, followed by the Orthodox religion **(Table 2).**

**Table 2:** Mother’s background characteristics for trends and determinants of neonatal mortality in rural Ethiopia, EDHS, 2011, 2016 and 2019.

| Variable | Category | Frequency | Percentage |
| --- | --- | --- | --- |
| Number of living children | <3 | 7132 | 31.3 |
|  | >=3 | 15623 | 68.7 |
| Maternal Age in years | <=24 years | 5599 | 24.6 |
|  | 25-34 years | 11599 | 51.0 |
|  | > 35-49 years | 5557 | 24.4 |
| Wealth index | Poor | 14050 | 61.7 |
|  | Middle | 4022 | 17.7 |
|  | Rich | 4683 | 20.6 |
| Number of antenatal visits during pregnancy | Not optimal | 11408 | 50.1 |
|  | Optimal | 3495 | 15.4 |
|  | Unknown | 7852 | 34.5 |
| Marital Status | Never married | 93 | 0.4 |
|  | Married/Living together | 21443 | 94.2 |
|  | Widowed/Divorced | 976 | 4.3 |
|  | Not living together | 243 | 1.1 |
| Maternal educational level | No education | 16387 | 72.0 |
|  | Primary | 5580 | 24.5 |
|  | Secondary | 616 | 2.7 |
|  | Higher | 172 | 0.8 |
| Religion | Orthodox | 6178 | 27.2 |
|  | Catholic | 181 | 0.8 |
|  | Protestant | 4429 | 19.5 |
|  | Muslim | 11487 | 50.5 |
|  | Traditional | 252 | 1.1 |
|  | Other | 228 | 1.0 |
| **Total** |  | **22755** | **100** |

### Household and Health service Related Characteristics

In this study of households’ access to facility characteristics, we found that 15715 (69.1%) of mothers use unprotected sources of drinking water, and 12989 (57.1%) do not access toilet facilities, while the remaining 7040 (30.9%) and 9766 (42.9%) of rural mothers access protected drinking water and toilet facilities, respectively. The summary table also reveals that more than half of the surveyed population, 69.6%, did not have access to television and radio. More than 90% of these mothers use wood as a fuel for cooking and other activities. Access to facilities such as protected sources of drinking water, utilization of toilet facilities, access to the media, and use of electricity for cooking and food preparation as a fuel has a likelihood to support mothers and childcare, implying a decrease in infant mortality **(Table 3).**

**Table 3:** Facility accessibility characteristics for trends and determinants of neonatal mortality in rural Ethiopia, EDHS,2011, 2016 and 2019.

| Variable | Category | Frequency | Percentage |
| --- | --- | --- | --- |
| Source of drinking water | Protected | 7040 | 30.9 |
|  | Unprotected | 15715 | 69.1 |
| Type of toilet facility | Use toilet facility | 9766 | 42.9 |
|  | No toilet facility | 12989 | 57.1 |
| Access to Media | No access to media | 15844 | 69.6 |
|  | With access to media | 6911 | 30.4 |
| Type of cooking fuel | Electricity | 67 | 0.3 |
|  | Gas | 18 | 0.1 |
|  | Charcoal | 403 | 1.8 |
|  | Wood | 20623 | 90.6 |
|  | Agricultural crop | 294 | 1.3 |
|  | Animal dung | 811 | 3.6 |
|  | Other | 539 | 2.4 |
| **Total** |  | 22755 | 100 |

### Neonatal and delivery related characteristics

As depicted in the table below, 51.4% of the children in the sample are male, while 48.6% are female. This indicates a slight imbalance in the sex distribution, with slightly more males than females. The distribution of neonatal mortality status in rural areas between the 2011, 2016, and 2019 EDHS data of this study disclose that 6.9% of the neonates in the sample died, while 93.1% survived. This provides insight into the mortality rate among neonates in the studied population. The status of the infant regarding the time she or he started breastfeeding was analyzed and found that 59.5% of the children were put in the breast immediately after birth, while 40.5% were not. This suggests that a significant portion of children did not receive immediate breastfeeding after birth. The distribution of child sizes at birth was also another variable of interest, which shows that 23.9% were classified as large, 31.9% as average, and 24.8% as smaller. Additionally, 19.4% have an unknown size. This indicates variability in the size of children at birth within the sample, with a notable proportion having an unknown size **(Table 4).**

**Table 4:**
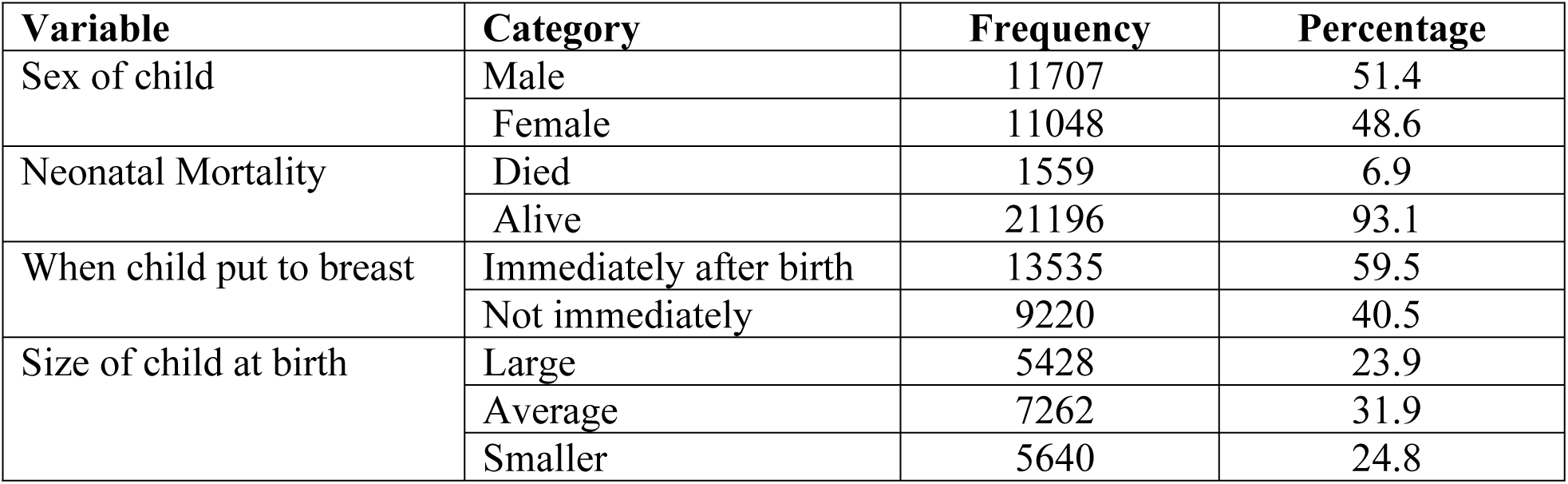
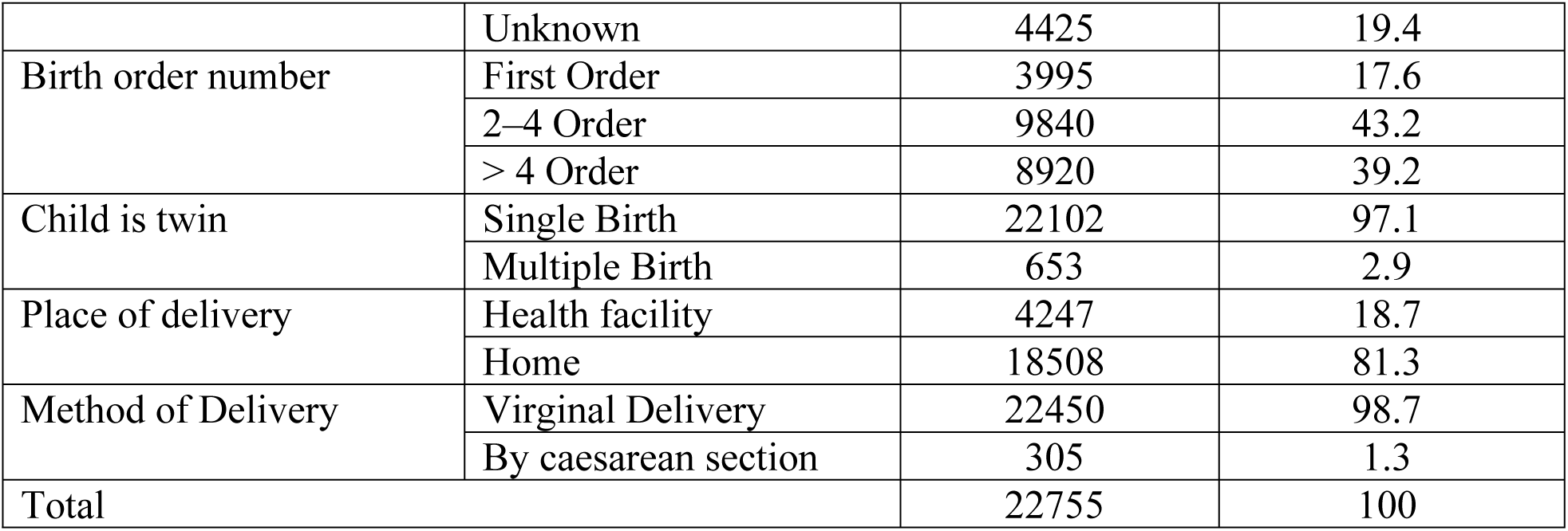
Neonatal and maternal related characteristics for trends and determinants of neonatal mortality in rural Ethiopia, EDHS, 2011, 2016 and 2019.

The majority of neonates in the study were reported being single births, 97.1%), and only 2.9% were multiple births (twins). This suggests that the vast majority of births included in the study involved single births, with only a small percentage being multiple births. Place of the neonate delivery was also the study variable implying that 81.3% of the deliveries occurred at home, and the remaining 18.7% of the deliveries took place in a health facility. This distribution shows that a significant majority of deliveries in the rural areas of Ethiopia were conducted at home rather than in a health facility. Substantial types of deliveries encompassed in the study were vaginal deliveries, 98.7%. Only 1.3% of the deliveries were by cesarean section. This indicates that the vast majority of deliveries in the rural areas of Ethiopia were vaginal deliveries, with only a small percentage requiring a cesarean section **(Table 5).**

**Table 5:** Trends of neonatal live status across years for trends and determinants of neonatal mortality in rural Ethiopia, EDHS, 2011, 2016 and 2019.

| Ethiopian<br>DHS Year | Neonate's live status |  |  |  | Total |
| --- | --- | --- | --- | --- | --- |
|  | Alive |  | Died |  |  |
|  | Frequency | Percentage | Frequency | Percentage |  |
| 2011 | 8,939 | 92.51 | 724 | 7.49 | 9,663 |
| 2016 | 8,099 | 93.45 | 568 | 6.55 | 8,667 |
| 2019 | 4,158 | 93.97 | 267 | 6.03 | 4,425 |
| Total | 21,196 | 93.15 | 1,559 | 6.85 | 22,755 |

### Trends of neonatal live status in rural areas

There is a notable EDHS-year disparity in neonatal mortality status. The DHS 2011 encountered the highest number of neonatal deaths, 724 (7.5%), followed by DHS 2016 (568 (6.6%)). In the 2019 DHS data, the slightest numbers of neonatal deaths were recorded. This implies that the risk of neonatal mortality in rural Ethiopia is decreasing. This decrease in neonatal mortality rates across the DHS years in rural Ethiopia is displayed in the figure below **(Table 5).**

This table presents data on total mortality in different regions of Ethiopia across 2011, 2016, and 2019 years as recorded in the Ethiopian DHS (Demographic and Health Survey). The total mortality rate in Tigray decreased over the years from 2011 to 2019. In 2011, 6.7% of children died, while in 2019, it decreased to 3.6%. The total mortality rate in Afar also decreased over the years, from 8% in 2011 to 4.4% in 2019. Similar trends are observed in other regions, with a decrease in total mortality rates from 2011 to 2019, indicating potential improvements in child health and healthcare services **(Table 6).**

**Table 6:** Trends of neonatal mortality by region in rural Ethiopia, EDHS, 2011, 2016 and 2019.

| Region | Ethiopian DHS Year |  |  |  |  |  | Total Mortality |  |
| --- | --- | --- | --- | --- | --- | --- | --- | --- |
|  | 2011 |  | 2016 |  | 2019 |  | Frequency (N) | Percentage (%) |
|  | N | % | N | % | N | % |  |  |
| Tigray | 71 | 6.7 | 54 | 6.3 | 25 | 6.6 | 150 | 3.6 |
| Afar | 79 | 8 | 71 | 7.3 | 45 | 8.2 | 195 | 4.4 |
| Amhara | 84 | 7 | 48 | 5.4 | 21 | 4.6 | 153 | 3.3 |
| Oromiya | 131 | 8.2 | 106 | 7 | 45 | 7.1 | 282 | 4.1 |
| Somali | 53 | 7 | 87 | 7.2 | 40 | 7.6 | 180 | 4 |
| B.Gumuz | 63 | 6.7 | 47 | 5.7 | 22 | 4.7 | 132 | 3.3 |
| SNNP | 117 | 7.7 | 63 | 5.3 | 25 | 4.2 | 205 | 3.4 |
| Gambela | 44 | 5.9 | 43 | 8 | 23 | 6.5 | 110 | 3.8 |
| Harari | 36 | 8.3 | 32 | 7.9 | 12 | 4.9 | 80 | 4.2 |
| Dire Dawa | 46 | 10.9 | 17 | 6 | 9 | 4.1 | 72 | 4.4 |
| Total | 724 | 7.5% | 568 | 6.6 | 267 | 6 | 1559 | 6.9 |

Total mortality rates vary between regions and across survey years. In 2019, as an illustration, the regions with the lowest mortality rates include Gambela (3.8%) and Tigray (3.6%), while the highest mortality rates are observed in Dire Dawa (4.4%) and Somali (4%).

While the general trend indicates a decline, there are regional variations in the pace and magnitude of this decline. Some regions show more significant reductions in neonatal mortality rates compared to others. For example, regions like Amhara, Benishangul Gumuz, SNNPR, and Dire Dawa exhibit consistent decreases in mortality rates over the years, indicating successful interventions or improvements in healthcare infrastructure and services. On the other hand, regions like Afar, Somali, Gambella, and slightly Oromiya may show less pronounced declines or even fluctuations in neonatal mortality rates, suggesting potential challenges or disparities in healthcare access or quality. This comparison provides insights into how each region fares compared to the national average.

### Factors Associated Neonatal Mortality in Rural Ethiopia

Displayed on this table are the logistic regression results pertaining to neonatal mortality, with the categorization of 1 for death and 0 for survival, and its relationship to various independent variables. Notably, the results reveal a significant negative correlation between Ethiopian DHS years and the probability of neonatal mortality.

A comparison of data, from Ethiopian Demographic and Health Surveys (EDHS) in 2011, 2016 and 2019, shows that neonatal mortality is decreasing over time. Neonates surveyed in 2016 were 13% less likely to die compared to those in 2011, while newborns in 2019 were 34% less likely to die than those in 2011. The age of the mother is also a significant factor. Neonates born to mothers aged 25-34 years were 14% less likely to die than those born to mothers aged less than 25 years. No significant association was found for mothers aged 35-49 years. The study found that neonatal mortality is negatively correlated with the rich wealth index. The odds ratio was 0.84 (p = 0.00), meaning that a child born to a wealthy mother has a 16% lower chance of dying compared to a child born to a poor mother. Families with three or more living children have 88% lower odds of neonatal mortality than families with less than three living children, according to the study. The adjusted odds ratio was 0.12.

The study examined the impact of birth order on the likelihood of neonatal death, dividing it into three categories: first order, second to fourth order, and more than fourth order. The results revealed that both second to fourth-order and above-fourth-order were statistically significant predictors of neonatal death, with infants after the first birth order exhibiting a significantly higher likelihood of death compared to the first birth order. Specifically, neonates in the 2-4 birth order have approximately 180% times higher odds of death when compared to the first birth order, holding constant the effect of other variables in the model. A study found that neonates born at higher birth orders have significantly higher odds of death. Infants from multiple deliveries are more likely to die compared to singleton deliveries. The gender of a newborn was found to have a significant effect on their likelihood of survival. Female infants were 20% less likely to die compared to male infants (OR = 0.80, P = 0.00). The time of breastfeeding after birth was also analyzed, and it was found that immediate breastfeeding was significantly associated with a 99% decrease in the odds of death compared to delayed breastfeeding at birth (OR = 0.01, P = 0.00), while other factors were held constant. These findings indicate a strong relationship between neonatal mortality and delayed breastfeeding after birth in rural areas among the population surveyed. Furthermore, the odds of neonatal death were 35% lower for mothers who utilized optimal antenatal care service visits compared to those who did not attend these visits (OR = 0.65, P = 0.00).

The model includes the size of the baby at birth, and it shows that babies born with a medium size have a significant negative association (OR = 0.75, P = 0.00), while babies born with a smaller size do not have a significant association. This means that babies born with a medium size are 25% more likely to die than babies born with a large size.

The model’s McFadden’s R square value is 0.2098, indicating that the independent variables included in the model account for approximately 21% of the variance in neonatal mortality. In addition, the model has a relatively good fit to the observed data, as the log-likelihood is - 5670.54. The link test with a p-value of 0.1369 suggests that there is no strong evidence of a lack of fit, and the overall model fits statistically significantly. **(Table 7).**

**Table 7:** Multiple logistic regression results predicting neonatal mortality in rural Ethiopia, EDHS, 2011, 2016 and 2019.

| Neonatal Mortality | B(95% CI) | SE | Wald | AOR(95% CI) | P-value |
| --- | --- | --- | --- | --- | --- |
| (Constant) | -1.52(-1.75,-1.30) | 0.12 | -13.14 | 0.20((0.16,0.24) | 0.00 |
| EDHS Year (2016) | **-0.16(-0.28,-0.04)** | **0.06** | **-2.55** | **0.87(0.77,0.98) \*** | **0.01** |
| EDHS Year (2019) | **-0.43(-0.60,-0.25)** | **0.09** | **-4.70** | **0.66(0.56,0.79) \*** | **0.00** |
| Maternal Age (25-34 yrs.) | -0.16(-0.31,0.00) | 0.08 | -1.93 | 0.86(0.74,1.01) | 0.05 |
| Maternal Age (35-49 yrs.) | 0.08(-0.13,0.29) | 0.11 | 0.74 | 1.09(0.88,1.34) | 0.46 |
| Wealth Index (Middle) | -0.09(-0.25,0.07) | 0.08 | -1.13 | 0.95(0.82,1.11) | 0.26 |
| Wealth Index (Rich) | **-0.23(-0.39,-0.07)** | **0.08** | **-2.86** | **0.84(0.73,0.97) \*** | **0.00** |
| No. of Living Child ($\geq 3$) | **-2.11(-2.29,-1.93)** | **0.09** | **-22.83** | **0.12(0.10,0.15) \*** | **0.00** |
| Birth Order (2–4 Order) | **1.03(0.85,1.21)** | **0.09** | **11.38** | **2.80(2.34,3.34) \*** | **0.00** |
| Birth Order (>4 Order) | **1.86(1.60,2.11)** | **0.13** | **14.49** | **6.39(4.97,8.21) \*** | **0.00** |
| Twins (Multiple) | **1.35(1.14,1.55)** | **0.10** | **13.00** | **3.83(3.13,4.69) \*** | **0.00** |
| Child Sex (Female) | **-0.22(-0.34,-0.11)** | **0.06** | **-3.96** | **0.80(0.72,0.89) \*** | **0.00** |
| BF Time (Instantly) | **-4.40(-4.92,-3.89)** | **0.26** | **-16.78** | **0.01(0.01,0.02) \*** | **0.00** |
| No. of ANC (Optimal) | **-0.43(-0.64,-0.21)** | **0.11** | **-3.80** | **0.66(0.53,0.82) \*** | **0.00** |
| No. of ANC (Unknown) | **0.28(0.15,0.41)** | **0.06** | **4.33** | **1.32(1.17,1.50) \*** | **0.00** |
| Child Size at Birth (Average) | **-0.28(-0.43,-0.14)** | **0.07** | **-3.80** | **0.75(0.65,0.87) \*** | **0.00** |
| Child Size at Birth (Smaller) | -0.13((-0.29,0.02) | 0.08 | -1.72 | 0.87(0.74,1.01) | 0.09 |

Note: B = unstandardized coefficient, SE = standard error, Wald = Wald statistic, OR = odds ratio. \* P < 0.05.

## 4. Discussion

This study was conducted to examine trends and determinants of neonatal mortality in rural areas of Ethiopia by using the Ethiopian demographic health survey in 2011, 2016, and 2019 In rural Ethiopia, trends of neonatal mortality from 2011 to 2016 decreased from 7.5% to 6.6%, unlike the very slow decline from 2016 to 2019, which was only 6%. This might be because Ethiopia has implemented ANC, postnatal care, immunization during and after pregnancy, the birth corner, basic life support, and birth attendance skills during the period before 2016[30]. This doesn’t mean that Ethiopia is not working with the listed strategies from 2016 to 2019 and till now, but the level of focus is different. This means the implementation of the strategies in rural areas of Ethiopia might be decreased due to give-back services, which may increase neonatal mortality.

In this study, the trends of neonatal mortality in most regions of Ethiopia increased, especially from 2016 to 2019. This finding goes together with the national neonatal mortality data, which shows the incidence of neonatal mortality from 29 to 33 per 1000 live births[5]. The possible justification could be due to insufficient health extension workers at health posts to deliver the package of essential health services, and the quality of care they provide is suboptimal due to different factors such as budget constraints, unlike the previous period[31].

From the multiple logistic regression, neonates included in the 2016 EDHS are 14.8% (CI: 0.283–0.037) less likely to die compared to those included in the 2011 EDHS. Here, the analysis shows that time has its effect on neonatal mortality, which could be defined by different factors such as the modernization and emergence of neonatal devices, scientific evidence, and the use of neonatal-trained human resources that interns used to give more qualified neonatal care that decreased the death rate. However, the probability of death among neonates included in the 2019 EDHS was 34.6% (CI: 0.602, 0.248) lower than neonates included in the 2016 EDHS. This finding shows that the probability of death over the recent period is lower, but there is a significant difference in each comparison EDHS period. Institutional level readiness differences during each period, the availability of neonatal intensive care units in most hospitals, and cultural issues that claimed that neonatal deaths were considered a secret and nothing or some bad punishment from the creator of the people that collectively affect the care of neonates that ends with death[32].This justification works since the attitude, infrastructure, and multiple issues have been fixed from ancient to recent times. However, there is still a breakup to sustain the decrease in neonatal mortality from 2011 to 2016 and 2019 EDHS.

This study identify that being born from wealth index (rich) family decreases the probability of neonatal death by 20.8% (CI:0.394,0.073) compared to their counter parts. This finding is similar with studies conducted in South Asia[33],Ghana[34]and in Ethiopia[35].The possible justification could be that factors contributing to the high neonatal mortality rate in poor families might be the lack of access to quality healthcare, maternal malnutrition, which may cause complications, inadequate sanitation and living conditions, which may increase maternal infection, and limited education and awareness about the dangers of pregnancy.

Neonates from a family with a living child of three or more have 88% (CI: 10, 0.15) less chance of death than those found with less than three. The possible justification could be due to limited resources; families with fewer members may have limited financial resources and access to healthcare services; mothers with lower family sizes may be younger; and young mothers often face increased risks during pregnancy and childbirth, which could boost neonatal mortality. However, this finding from this study is contrary to the conclusion reported by the study conducted in Ethiopia using the 2019 EDHS[36].

Neonates with birth order 2–4 and >4 order have 2.80(CI: 2.34, 3.34) and 6.39(CI: 4.97, 8.21) times risk of death compared with birth order 2 and less. This finding is almost similar with the study conducted in Ethiopia[36,37].The possible justification for this finding could be due to increased maternal age, which can be accompanied by different neonatal complications secondary to maternal health, including prematurity.

The odds of twin /multiple pregnancy neonatal death is 3.8times(CI:3.13,4.69)) higher than singleton pregnancy .The finding of this study is in line with previously conducted study in Ethiopia[38].This can be due to the fact that twin pregnancy is highly risk for preterm birth, small for gestational age, low birth weight and intrapartum anoxia which individually or collectively causes for neonatal mortality[39].

Based on the finding of this study being femaleness have 20 %(CI: 0.72, 0.89) less likely to died compared with male neonates. This conclusion agreed with different studies [7,29]. The possible justification for this find might not be biological sound despite different methodological, birth prevalence difference between the two sexes.

The preventive effects of early breast feeding initiation among neonates is 99 %(CI: 0.01, 0.02) compared with neonates with delayed breastfeeding initiation. This finding agreed with different previous studies [29,38,40].This might be due to multiple uses of first vaccine (breast milk) which protects the newborn from acquiring infection, decrease complications and reduces newborn mortality.

In this study, optimal ant-natal care has a positive association with neonatal mortality, which decreases the risk by 34% (CI: 0.53, 0.82), whereas unknown or no ant-natal care increases the probability of neonatal death by 1.32 times (CI: 1.17, 1.50) compared with neonates born to mothers having regular ant-natal follow-up care. This finding is comparable with the previous studies[30,35,36].This finding is supported by scientific evidence in which antenatal care is intended to ensure that a woman has a safe pregnancy and allows screening of preeclampsia, fetal abnormalities, and other prevention strategies to be incorporated. As a result, early identification and management will be applied, which can decrease neonatal mortality[41].

The probability of neonatal death from a probability family of average child size is 25% (CI: 0.65, 0.87) less than neonates from a large family size. This finding is also in line with previously conducted studies[5,30,36].This might be due to the larger the family, the less economical it may be to serve the family, especially the pregnant mother with her fetus, resulting in different perinatal complications and a poor neonatal outcome, especially in rural areas.

### Study Limitation

First, the results from the cross-sectional study only refer to one point in time. As a result it, poses an inability to make cause-and-effect analyses.

## 5. Conclusions

Regions such as Amhara, Benishangul Gumuz, SNNPR, and Dire Dawa have displayed consistent decreases in mortality rates, reflecting successful interventions or enhancements in healthcare systems and services.

Conversely, regions like Afar, Somali, Gambella, and to a lesser extent Oromia may exhibit less prominent declines or even fluctuations in neonatal mortality rates, suggesting potential challenges or disparities in healthcare accessibility or quality.

Additionally, factors such as maternal age, birth order, mothers’ wealth index, infant sex, birth size, presence of twins/multiple births, the number of antenatal care visits, and immediate breastfeeding after birth were found to be significantly associated with neonatal deaths.

## Abbreviations

ANC: Antenatal care
DHS: demographic health survey
EDHS: Ethiopia demographic health survey
EMDHS: Ethiopia mini demographic health survey
LMICs: Low and middle-income countries
MDG: Millennium Development Goal
NMR: Neonatal mortality rate
SDG: Sustainable Development Goals

## Data Availability

The datasets used and/or analyzed during the current study available from the corresponding author on reasonable request.

NA

## 6. Declarations

### Consent for publication

Not applicable.

### Competing interests

The authors declare that they have no competing interests.

### Funding

Not applicable. EDHS data used to conduct the study.

### Authors’ contributions

All authors contributed to the preparation of the manuscript. S.A. prepared the draft manuscript D.S. collected and conducted analysis then, D.B. and T.G. revised the analysis. M.B., M.G., G.T., N.B., Y.S., F.A., and D.T. revised the final draft of the paper. All authors read and approved the final manuscript.

### Ethical consideration

In order to utilize the data for research purposes, we obtained a formal grant from the EDHS team. We followed the ethical principle of not sharing any of the data with anyone.

## Acknowledgments

We would like to thank the EDHS team for providing us with the data to analyze and draw conclusions.

## Notes

### Competing Interest Statement

The authors have declared no competing interest.

### Clinical Trial

NA

### Clinical Protocols

NA

### Funding Statement

The author(s) received no specific funding for this work.

